# Disrupted-in-schizophrenia 1 (DISC1) protein aggregates in cerebrospinal fluid are elevated in first-episode psychosis patients

**DOI:** 10.1101/2023.04.17.23288687

**Authors:** Marlene Pils, Julia Rutsch, Feride Eren, Göran Engberg, Fredrik Piehl, Simon Cervenka, Carl Sellgren, Svenja Troßbach, Dieter Willbold, Sophie Erhardt, Oliver Bannach, Carsten Korth

## Abstract

The Disrupted-in-schizophrenia 1 (DISC1) protein is a key regulator at the intersection of signaling pathways relevant for adaptive behavior. It is prone to posttranslational changes such as misassembly and aggregation but the significance of such transformations for human mental illness has remained unclear.

Here we demonstrate that DISC1 protein aggregates are increased in CSF samples of patients with first episode psychosis (n=50) compared to healthy controls (n=47), as measured by the highly sensitive surface-based fluorescence intensity distribution analysis technology that enables single aggregate detection. The concentration was in the low femtomolar range. No correlations were found to symptom levels, but the difference was particularly significant in the subset of patients receiving the diagnoses “schizophrenia, unspecified” (DSM IV 295.9) or schizoaffective disorder (DSM IV 295.70) at 18-month follow-up.

The occurrence of protein aggregates *in vivo* in patients with psychotic disorders has not been previously reported. It underscores the significance of posttranslational modifications of proteins both as pathogenetic mechanisms and as potential diagnostic markers in these disorders.

## Main

Molecular markers for major psychiatric diseases, such as schizophrenia and recurrent affective disorders that can aid diagnostic workup and/or therapeutic monitoring are urgently needed. The current lack of such markers in parts reflects an incomplete understanding of underlying disease mechanisms. Initial hopes of genetic markers to fulfill such a role have not materialized; although schizophrenia shows a high degree of heritability, currently known common genetic variants together account for less than 10% of the variance in disease risk ^1^.

Proteins are the ultimate executors of cellular functions and are involved in signaling pathways governing adaptive behavior. They are also the most recognized molecular markers in medical routine diagnostics, usually detected by specific immunological assays. Posttranslationally modified proteins play a key role in neurodegenerative diseases, where the presumed causally responsible protein aggregates such as Aβ, tau, α-synuclein, or others can be measured in *post mortem* brain, *in vivo* using PET, in cerebrospinal fluid (CSF) and in blood.

The establishment of a role of posttranslationally modified proteins, and particularly the identification of candidate proteins, in major psychiatric diseases, is still in its infancy ^2,3^. The disrupted-in-schizophrenia 1 *(DISC1*) gene was identified as a familial gene in a large Scottish pedigree ^4^ but was subsequently not found to feature in larger studies of common gene variants ^1,5^. The protein, however, is subject to posttranslational modifications such as phosphorylation ^6^ and multimerization ^7^. Furthermore, it is highly aggregation-prone, both *in vitro* and *in vivo* ^7,8^. In *post mortem* brains, DISC1 aggregates have been identified biochemically in schizophrenia and recurrent affective disorders, but were not found in healthy controls or patients with classical neurodegenerative diseases ^7,9^. An animal model displaying DISC1 aggregation showed dysregulated dopamine homeostasis ^10^, consistent with a role for DISC1 aggregates in non-adaptive behavior.

We set out to identify DISC1 aggregates in patients with first episode psychosis using a highly sensitive surface-based fluorescence intensity distribution analysis (sFIDA). Previously, we introduced sFIDA as a protein oligomer-specific quantification method for counting single proteinaceous particles. sFIDA features a combination of an ELISA-like biochemical setup and microscopy-based readout with sub-femtomolar sensitivity ^11-14^. In the hereby for DISC1 perotein aggregates adapted sFIDA assay, samples are incubated on a capture antibody-coated glass surface and both monomeric and aggregated DISC1 species are immobilized. However, since sFIDA uses the same or overlapping epitopes for capturing and detecting, monomeric species are not detected due to the epitope already being blocked by the capture antibody. Specificity is further increased using two differently labeled detection probes targeting the same or overlapping epitopes (Supplementary Fig. 1). After imaging the glass surface by dual-color total internal reflection fluorescence microscopy (TIRFM) only those aggregates are counted that yield signals in both fluorescence channels (co-localization). To reduce background noise, an intensity cutoff is applied and only signals above the cutoff were evaluated (sFIDA readout).

The main aim of our study was to use sFIDA to compare quantitative DISC1 protein aggregates in CSF between first episode psychosis patients (FEP) vs. healthy controls to explore the presence and the potential diagnostic significance of DISC1 aggregate detection for clinical use. A second aim was to compare DISC1 aggregate levels to the positive and negative syndrome scale (PANSS), as well as other cognitive dimensions quantified by established psychological tests. Finally, DISC1 aggregates levels were compared between subsets of patients depending on their DSM-IV diagnosis at follow-up. to explore the use of DISC1 as a potential biomarker for clinical progression. Assay development and analytical validation (online methods) were performed using artificial DISC1 aggregates, which were either DISC1-coated SiNaPs or synthetic DISC1 (598-785) aggregates ^15^ spiked into buffer showing femtomolar detection limits (6.08 fM LoD and 9.54 fM LLoQ) and low intra-assay variability (DISC1 SiNaPs 15.8%, synthetic DISC1 aggregates 7.9%, Supplementary Fig. 2), high assay comparability (differences less than 10%, Supplementary Fig. 3) and complete selectivity (∼100%, Supplementary Fig. 4).

DISC1-coated SiNaPs, synthetic DISC1 aggregates, and a set of 97 CSF samples from first-episode psychosis patients (FEP, n=50, 31 male, 19 females) and healthy controls (HC, n=47, 20 male, 27 females) enrolled in the Karolinska Schizophrenia Project were subjected to sFIDA analysis (Fig. 1). Patient and control samples were mixed and analyzed blinded to group allocation. Patients that upon followup did not receive a clinical diagnosis were excluded from the overall analysis. To account for measurement bias on multiple plates, samples were normalized using an individual plate specific normalization factors based on the readouts of the HC group. Detailed information on the calculation and used normalization factors see online methods and Supplementary Tab. 1. Since normalized data showed non-normal distribution (Supplementary Tab. 2), we used two-sided Mann-Whitney U test, which revealed that concentrations of aggregated DISC1 species were significantly increased in CSF samples of FEP patients compared to HC (p-value 0.026, Fig. 1a). Representative TIRFM images of DISC1 SiNaPs, synthetic DISC1 aggregates, blank control and a human CSF sample from FEP patient are shown in Fig. 1b.

**Fig. 1.**
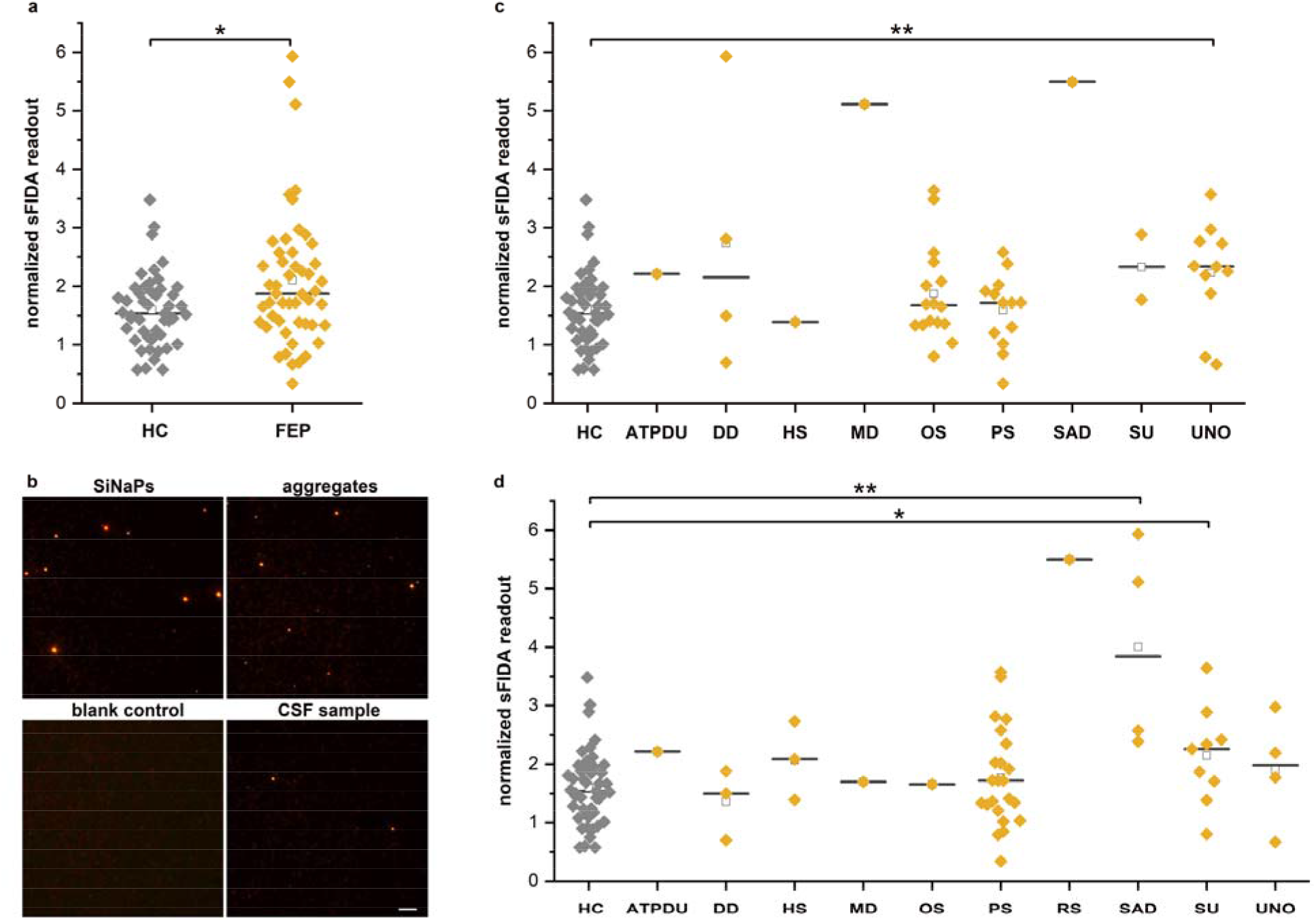
Quantitation of DISC1 aggregates in CSF samples of first-episode psychosis patients and healthy controls using sFIDA technology. **(a)** Scatter plot of normalized sFIDA readouts for healthy controls (HC, n=47) and first-episode psychosis patients (FEP, n=50). Statistical analysis was performed using two-sided Mann-Whitney U test (confidence interval of 0.05), and significantly elevated DISC1 levels in FEP patient samples were determined, indicated by a p-value of 0.027. **(b)** Representative TIRFM images as a composition of channel 0 and channel 1 (with an excitation: 633 nm, emission: 705 nm (channel 0) and an excitation: 488 nm, emission: 525 nm (channel 1), both with an exposure time of 500 ms and gain of 800) of DISC1 SiNaPs (160 fM), synthetic DISC1 aggregates (32 fM, sub-unit concentration), blank control (blocking solution only) and human CSF sample (FEP patient sample). Scale bar: 20 μm. Samples were divided according to their diagnosis at baseline **(c)** or at follow-up **(d)** and examined for significant differences from HC samples using one-sided Mann Whitney U test (confidence interval of 0.05). Using the baseline diagnosis for grouping, significantly higher DISC1 levels were determined for UNO (p-value 0.005) compared to the healthy control group. In contrast, using the follow-up diagnosis for grouping revealed significantly higher DISC1 levels for SAD (p-value 0.001) and SU (p-value 0.024) compared to the healthy control group. The line indicates the median and the square indicates the mean. Significant differences between cohorts were signed with * (*p=0.01-0.05; **p=0.001-0.01). *HC* healthy control, *FEP* first-episode psychosis patients, *ATPDU* Acute, and transient psychotic disorders, unspecified (all diagnoses are DSM IV 298.8), *DD* Delusional Disorders (297.1), *HS* hebephrenic schizophrenia (295.10), *MD* Major depressive disorder (296.21), *OS* Other Schizophrenia (295.40), *PS* Paranoid schizophrenia (295.30), *RS* Residual Schizophrenia (295.60), *SAD* Schizoaffective disorder (295.70), *SU* Schizophrenia, unspecified (295.9), *U*NO Unspecified nonorganic psychosis (298.9).

Sex, BMI, and nicotine usage were not significantly different between healthy controls and FEP patients (demographic information Supplementary Tab. 3). Mean age (mean ± s.e.m) for healthy controls was 26.9 ± 0.8 years and for FEP patients 30.1 ± 1.2 years. No significant correlations were evident between CSF DISC1 aggregate levels and age, BMI, sex, smoking status, or medication (Supplementary Tab. 4 and Tab. 5).

We then looked at correlations of DISC1 protein aggregate levels with PANSS scores, Global Assessment of Functioning Scale, Clinical Global Impression Scale, as well as cognitive tests from the MATRICS battery but no significant result was obtained after false discovery rate correction (see Supplementary Tab.8).

When we separated different subsets of patients, based on the clinical diagnosis at follow-up, assuming that DISC1 aggregate levels might be more elevated in some subsets rather than in others. A one-sided Mann Whitney U test (confidence interval 0.05) was used to reveal differences between the respective subsets and healthy controls (Fig. 1d, Supplementary Tab. 7). Significantly higher CSF levels of aggregated DISC1 compared to controls were observed in schizoaffective disorder (SAD, DSM IV 295.70; p-value 0.001) and schizophrenia, unspecified (SU, DSM IV 295.9; p-value 0.024). The cases of SAD were in low numbers and thus making the present results preliminary. Moreover, the schizophrenia subtypes have shown to have low diagnostic reliability and validity, resulting in their removal in DSM V. Nevertheless, the present results suggest that DISC1 aggregate-positive cases may be clinically rather atypical and could overlap with symptoms from affective disorders. Supporting DISC1 aggregate positive cases as a possible diagnostic category not matching current clinical boundaries is also the absence of solid correlations to symptom measures. Interestingly, phenotypic variability spanning from schizophrenia to affective disorders has also been reported in the Scottish pedigree with DISC1 mutations where the *DISC1* gene was discovered ^4^. However, the low number of SAD cases limits possibilities to draw robust conclusions from the present data set.

It is noteworthy that from the initial diagnosis at baseline, only one clinical subset of patients featured significantly elevated DISC1 aggregate levels (unspecified nonorganic psychosis, UNO, Fig. 1c, Supplementary Tab. 6), but that in the more solid clinical diagnosis after 18 months, clinical subsets SAD and SU turned out to be significantly associated with elevated DISC1 aggregate levels. This was mainly due to some individuals that at baseline were diagnosed as “other schizophrenia” (OS, DRM IV 295.40) who upon follow-up instead fulfilled criteria for paranoid schizophrenia (PS, 295.30, seven cases) and unspecified schizophrenia (SU, DSM IV 295.9, five cases), and schizoaffective disorders (SAD, DSM IV 295.70, 1 case).

For 21 FEP patients (11 male, 10 female) and 15 HC (3 male, 12 females) in the present sample, CSF samples were obtained again after 18 months, allowing us to investigate longitudinal differences. However, for both HC (p-value 0.074) and FEP patients (p-value 0.487), no significant changes in DISC1 aggregate levels were detected by Wilcoxon rank test (Supplementary Fig. 5), indicating that DISC1 aggregate levels seem to be stable over the medium term, in turn indicating a more stable phenotype.

In summary, we here for the first time demonstrate that DISC1 protein aggregates can be detected CSF and that levels are increased in first episode psychosis patients. Increases were seen specifically in the subsets of unspecified schizophrenia or schizoaffective disorder. The existence of a subset of psychosis patients featuring high DISC1 aggregate levels, previously termed DISC1opathies ^16^, strengthens the notion of this brain proteinopathy representing a novel pathogenic mechanism independent of familial or common genetic traits, and highlights it as a candidate biomarker for phenotypic characterization in FEP.

## Data Availability

The data that support the findings of this study are available on request from the corresponding author. The data are not publicly available due to privacy or ethical restrictions. For image data analysis, we used the sFIDAta software tool, which can be made available upon request from the corresponding author

## Data availability

The data that support the findings of this study are available on request from the corresponding author. The data are not publicly available due to privacy or ethical restrictions

## Code availability

For image data analysis, we used the sFIDAta software tool, which can be made available upon request from the corresponding author

## Acknowledgements

We thank Volker Nischwitz (Central Institute for Engineering, Electronics and Analytics, Analytics (ZEA-3), Forschungszentrum Jülich, 52428, Jülich, Germany) for determination of silicon concentration of the used DISC1 SiNaPs by ICP-MS. C.K. was supported by a grant from the DFG 1679/14-1. The study was further supported by the Swedish Research Council (Grant No. 523-2014-3467 (SC), Erling Persson Foundation (CMS), 2021-02251_VR (SE), 2019-01452_VR (GE)), Karolinska Institutet and Stockholm County Council (20160328, 20180487 [SC], (20190175) [SE], 20190447 [CMS]). Development of the sFIDA technology was supported by the programs “Biomarkers Across Neurodegenerative Diseases I + II” of The Alzheimer’s Association, Alzheimer’s Research UK and the Weston Brain Institute (11084 and BAND-19-614337). We are also grateful for support from The Michael J. Fox Foundation for Parkinson’s Research (14977, 009889), from the ALS Association and from the Packard Center (19-SI-476)

## Contributions

CK conceived the project. MP, ST and JR developed the assay. MP designed and performed the experiments and validation studies and analyzed the sFIDA data. CSM and SC coordinated recruitment and clinical characterization of the patients. F.P. coordinated and performed the lumbar punctures. MP wrote the manuscript in consultation with OB, SE, FE and CK. OB, SE, GE, SC, CS, DW and CK supervised the project. All authors discussed the results and provided critical feedback.

## References

1. Trubetskoy V, et al. Nature 604, 502–508 (2022).

2. Bradshaw, N.J. & Korth, C.. Mol Psychiatry 24, 936–951 (2019).

3. Hui KK, Endo R, Sawa A & M., T.. Biol Psychiatry 91, 335–345 (2022).

4. Millar, J.K., et al. Hum Mol Genet 9, 1415–1423 (2000).

5. Mathieson, I., Munafo, M.R. & Flint, J.. Molecular psychiatry 16, 1–8 (2011).

6. Ishizuka, K., et al. Nature (2011).

7. Leliveld, S.R., et al. The Journal of neuroscience 28, 3839–3845 (2008).

8. Cukkemane A, et al. Transl Psychiatry 11, 639 (2021).

9. Ottis, P., et al. Biol Psychiatry 70, 604–610 (2011).

10. Trossbach, S.V., et al. Mol Psychiatry 21, 1561–1572 (2016).

11. Kulawik A, Heise H, Zafiu C, Willbold D & O., B. FEBS Lett. 592, 516–534 (2018).

12. Herrmann, Y., et al. Clin Biochem 50(4-5):244–247, 244-247 (2017).

13. Kass B, et al. Cell Rep Med 3(2022).

14. Blömeke L, et al. NPJ Parkinsons Dis. 8(1):68, 68 (2022).

15. Leliveld, S.R., et al. Biochemistry 48, 7746–7755 (2009).

16. Korth, C.. Prion 6, 134–141 (2012).

