## Supplemental Methods Figures and Tables for "Disrupted-in-schizophrenia 1 (DISC1) protein aggregates in cerebrospinal fluid are elevated in first-episode psychosis patients"

### Supplemental Material

### Online Methods

#### Synthesis of silica nanoparticles coated with DISC738-778

To translate sFIDA readouts into molar aggregate concentration we introduced a silica nanoparticle-based calibration standard (SiNaPs) [1,2]. In the present study, we used SiNaPs coated with synthetic DISC1 peptide 738-778 (EDKRKTPLKVLEEWKTHLIPSLHCAGGEQKEESYILSAEL-Cysteamine-SH, peptides and elephants GmbH, Henningsdorf, Germany), the epitope of monoclonal antibody 14F2 ([3] see below), for assay development and as assay control. According to the previously published protocol [2], SiNaPs were synthesized via Stöber process and silica surface was functionalized with 3-aminopropyl(triethoxysilane) (APTES, Sigma-Aldrich, St. Louis, USA). Afterwards, the carboxy groups of maleimido hexanoic acid (MIHA, abcr GmbH, Karlsruhe, Germany) were activated by 1-ethyl-3-(3-dimethylaminopropyl)carbodiimide (EDC, Sigma-Aldrich, St. Louis, USA) and N-hydroxysuccinimid (NHS, Sigma-Aldrich, St. Louis, USA) and were coupled covalently to the introduced primary amines on silica surface. Maleimide groups and 14F2 peptides were crosslinked by using C-terminal functionalized DISC1 peptide. Finally, the molar SiNaPs concentration was calculated based on silicon concentration determined by inductively coupled plasma-mass spectrometry (ICP-MS) and physical characteristics like size, density as well as shape determined by transmission electron microscopy (TEM).

#### Synthetic DISC1 aggregates

In addition to SiNaPs, synthetic DISC1 aggregates were prepared for assay development and analytical validation. To this end, the human DISC1 protein fragment including codon-optimized amino acid residues 598-785 (DISC1 598-785) was cloned and expressed as described previously [4], except that the protein was refolded while on the Ni-NTA column (Qiagen GmbH Germany, Hilden, Germany) by sequentially diluting the 8 M urea extraction buffer by extraction buffer without urea (5 volumes). After elution with 100 mM imidazol-containing elution buffer, DISC1

(598-785) was dialyzed against PBS 10 mM DTT and sonicated in a Misonix S4000MPX sonicator (Qsonica L.L.C, Newtown, US) for 2.5 min at 93.9 kJ. This preparation was found to be stable in aggregation size over weeks. The majority of the multimers (99%) were determined to possess a hydrodynamic diameter of 41 nm by dynamic light scattering, corresponding to a 421mer, i.e. a large multimer of DISC1 (598-785).

**Assay antibody 14F2**

For the anti-human DISC1 monoclonal IgG antibody 14F2 (747-768), mice were immunized with recombinant C-terminal human DISC1 fragment (598-785). Hybridoma were generated, the antibody was harvested from FCS-free supernatant [3], dialyzed against PBS and concentrated (Amicon filter tubes, MWCO:10, 2700x g for 30 min at 4 °C, Merck, Darmstadt, Germany) to a stock concentration of 2.4 mg/ml.

In the sFIDA assay, 14F2 was used for capturing and detection of DISC1 aggregates. For microscopic detection of DISC1 aggregates, 14F2 was labelled according to manufactures' instructions with CF633 and CF488A dye (Biotium, Inc., Fremont, USA), respectively. In carbonate buffer, the succinimidyl ester group of the pre-activated dyes reacted covalently with the amines of the antibody. The fluorescent probes were purified using a polyacrylamide bead suspension (Bio-Gel P-30 Gel, Bio-Rad Laboratories, Inc., Hercules, USA) and were stored at 4 °C until further use. Antibody concentration and the degree of labelling was determined according to the manufacture's protocol.

**sFIDA assay protocol**

To reduce the risk of a microbial contamination, all in-house produced buffers and solutions were filtrated (VacuCap 60, 0,22 µm, Pall Corporation, Port Washington, USA) before usage and additionally, the whole sFIDA procedure was carried out under a clean bench to avoid dust and other contaminations. For all washing steps, a microplate washer (405 LS Microplate Washer, BioTek Instruments, Inc., Vermont, USA) was used.

For analytical as well as diagnostic validation, sFIDA assays were performed on 384-well microtiter plates (SensoPlate plus, Greiner Bio-One GmbH, Frickenhausen, Germany). As capture, the monoclonal anti-DISC1 antibody 14F2 (7.5 µg/ml in 0.1 M MES buffer (Carl Roth GmbH & Co.KG, Karlsruhe, Germany), pH 3.4) was immobilized on the glass well surface. After washing the wells five times with 80 µl TBS-T (1x TBS (Serva electrophoresis GmbH, Heidelberg, Germany) and 0.05% Tween20 (AppliChem GmbH, Darmstadt, Germany)) and five times with TBS, the unoccupied well surface was blocked with The Blocking Solution (Candor Bioscience GmbH, Wangen im Allgäu, Germany) for 1.5 h at RT. The plates were washed again with TBS-T and TBS as described above and 20 µl of DISC1 SiNaPs, synthetic DISC1 aggregates as well as of each human CSF samples (1:2 diluted in The Blocking Solution) were incubated for 2 h at RT. Afterwards, the wells were washed again with TBS (see above) and were incubated for 1 h at RT with the fluorescently labeled detection antibodies 14F2 CF688 and 14F2 CF488A (each 0.625 µg/ml) in TBS-T with 0.1% BSA (AppliChem GmbH, Darmstadt, Germany). To reduce background noise, unbound detection probes were removed by five washing steps (as described above) using TBS and finally, the washing buffer was replaced with TBS-ProClin (0.03% ProClin300, Sigma-Aldrich, St. Louis, USA) and the plates were sealed (SealPlate film, Sigma-Aldrich, St. Louis, USA) to avoid contaminations while measuring.

**Image Data Acquisition and data analysis**

Imaging of the well surface was performed using a total internal reflection microscope (TIRFM, Leica DMI600B, Leica microsystems GmbH, Wetzlar, Germany) as described previously by Kravchenko et al. [5] In total, 3.14% of the well surface were imaged in two different channels (channel 633: excitation: 635 nm, emission filter: 705/72 nm; channel 488: excitation: 488 nm, emission filter: 525/36 nm, exposure time: 500 ms, gain: 800). Images comprising 1000 x 1000 pixels and representing an area of 113.76 µm x 113.76 µm were analyzed using an in-house developed software that features automated artifact elimination and counting co-localized pixels. The sFIDA readout is referred to as the average number of pixels within an image that exceed a pre-defined cutoff value and are co-localized in both fluorescence channels. To this end, blank control-based cutoffs were defined as the signal intensity exceeding 0.05% for analytical validation studies, while the cutoff for the analysis of the whole dataset of human CSF samples was 0.005%. Due to fluctuations of the absolute fluorescence intensities for plate 1 of the diagnostic validation studies, for this plate MinMax filtering [2] with 5% was applied. Statistical analysis was performed using OriginPro2017 (OriginLab Corporation, Northampton, USA) and matlab2019b (The MathWorks, Inc., Natick, USA) software.

**Analytical validation**

Prior to measurement of the human CSF samples, the analytical performance of the developed sFIDA assay for DISC1 aggregates quantification was validated. As mentioned above, only DISC1 SiNaPs and synthetic DISC1 aggregates could be used for these studies, as human CSF samples were limited. For each study, the sFIDA surface was coated as described above. Afterwards, dilution series (1:5 in dilution buffer, quadruplicates) of both targets were applied according to individual study protocols and after incubation with detection probes, the assay surface was scanned via TIRFM. For analysis, a cutoff of 0.05% of blank control values was used to determine the number of co-localized DISC1 aggregates.

<u>Assay sensitivity and intra-assay variability</u>

First, we evaluated whether we could accurately detect and quantify DISC1 SiNaPs and synthetic DISC1 aggregates by investigating parameters such as Limit of Detection (LoD), Lower Limit of Quantification (LLoQ) and CV%. To this end, calibration standards and synthetic DISC1 aggregates were diluted in The Blocking Solution (1:5 dilution series) with concentrations ranging from 237 pM to 0.02 pM of DISC1 SiNaPs and 20 nM to 1.2 pM of synthetic DISC1 aggregates, respectively (Supplementary Fig. 2). Please note that the concentration of synthetic DISC1 aggregates refers to the used molar sub-unit concentration. Linear regression was performed with the sFIDA readouts weighted by 1/readout to calculate the calibration curve (matlab2019b software). For this calculation, only DISC1 SiNaPs concentrations were included that were in the linear measurement range and differed significantly from blank control (determined with one-sided Mann-Whitney U test with a confidence interval of 0.05%). Afterwards, LoD and LLoQ were calculated using Eq. 1a and Eq. 1b and translated into femtomolar concentration using determined calibration curve.

$$\text{(1a)} \quad \text{LoD [fM]} = \text{sFIDA readout (blank control)} + 3\sigma$$

$$\text{(1b)} \quad \text{LLoQ [fM]} = \text{sFIDA readout (blank control)} + 10\sigma$$

The intra-assay variability was defined by the CV% value, using mean and standard deviation of the quadruplicates for calculation while values below 20% are accepted and displays low variance between the replicates.

<u>Assay comparability</u>

To investigate inter-assay variability, dilution series of DISC1 SiNaPs and synthetic DISC1 aggregates were prepared and subjected on two different assay runs by the same operator with one-week time interval. In addition, to verify inter-machine variability, one of the prepared microtiter plate of inter-assay testing was measured on two different TIRF microscopes. Comparability was calculated by the percentage ratio of the observed and expected sFIDA readouts (Eq. 2) while values between 80-120% were accepted. For the illustration of the results, the measured sFIDA readouts of both measurements or for both TIRF microscopes were plotted against each other (Supplementary Fig. 3).

$$(2)\quad \text{assay comparability }[\%] = \frac{\text{observed sFIDA readouts}}{\text{expected sFIDA readouts}} \times 100\%$$

<u>Assay selectivity</u>

To determine assay selectivity, DISC1 SiNaPs (237 pM, molar particle concentration) and synthetic DISC1 aggregates (20 nM, molar sub-unit concentration) were applied on different sFIDA setups. Unspecific binding of DISC1 species to the blocking agent should be revealed by omitting the capture antibody (capture control, CC). In addition, autofluorescence (AF) from assay buffers was assayed by omitting the detection probe. Furthermore, the cross-reactivity of anti-Aβ-antibodies (CR) against the immobilized DISC1 species was also tested. For each assay control setup the ratio of observed and expected (reference, normal assay setup) values as well as the percentage amount of reduced signal was calculated according to Eq. 3a and Eq.3b.

$$(3a)\quad \frac{\text{O}}{\text{E}}[\%] = \frac{\text{observed readout assay control}}{\text{expected readout reference}} \times 100\%$$

$$(3b)\quad \text{signal reduction }[\%] = 100 - \text{O/E}$$

**Diagnostic validation**

After successful analytical validation of the developed sFIDA assay, we determine DISC1 aggregate levels in 169 human CSF samples using the sFIDA protocol described above. One sample was excluded from further analyses due to artificial images in all four replicates. In addition, after unblinding, we used only those CSF samples for further statistical analysis where clinical diagnoses were available both at the time of initial admission and re-assessment. Therefore, all further information and statistics refer to a total of 97 CSF samples including 50 samples from first-episode psychosis patients (FEP, n=50, 31 male, 19 females) and healthy controls (HC, n=47, 20 male, 27 females).

Patients

The present study was approved by the Stockholm Regional Ethics Committee (Dnr 2010/879-31/1). Prior to participating in the trial, all patients and HC completed a written informed consent form in line with the Helsinki Declaration. The Karolinska Schizophrenia Project (KaSP) recruited all participants in partnership with four psychiatric institutes in Stockholm, Sweden: PRIMA Vuxenpsykiatri, Södra Stockholms Psykiatri, Norra Stockholms Psykiatri, and Psykiatri Nordväst. The study's participants were recruited between March 2011 and March 2019. The presence of neurologic disorders or severe somatic sickness, a history of illicit substance addiction, and the existence neurodevelopmental abnormalities such as autism spectrum disorder were all exclusion factors for the study. Urine testing was used to assess drug usage. To rule out macroscopic brain abnormalities, magnetic resonance imaging (MRI) was employed. To assess clinical characteristics of the patients, the Global Assessment of Functioning (GAF; where symptom and functioning dimensions were assessed separately), the Positive and Negative Syndrome Scale (PANSS), Clinical Global Impression (CGI), Alcohol Use Disorders Identification Tests, and Drug Use Disorders Identification Tests were used. A clinical interview using the Diagnostic and Statistical Manual of Mental Disorders IV (DSM-IV), or a consensus diagnostic process was employed to establish the diagnosis. Patients were contacted for re-assessment after roughly 18 months, and the corresponding DSM-IV diagnosis were determined for 50 patients: paranoid schizophrenia (*PS*, DSM IV 295.30) (n=23), schizophrenia unspecified (*SU*, DSM IV 295.9) (n=9*),* other schizophrenia (*OS*, DSM IV 295.40) (n=1), hebephrenic schizophrenia (*HS*, DSM IV 295.10) (n=3), residual schizophrenia (*RS*, DSM IV 295.60) (n=1), delusional disorders (*DD*, DSM IV 297.1) (n=3), acute and transient psychotic disorder, unspecified (*ATPDU*, DSM IV 298.8) (n=1), unspecified nonorganic psychosis (*UNO*, DSM IV 298.9) (n=4), schizoaffective disorder (*SAD,*

DSM IV 295.70) (n=4), major-depressive disorder (*MD*, DSM IV 296.21) (n=1). Tobacco products were permitted to be used by patients and 13 of the patients (26%) used nicotine derivatives (smoking or snuff) [Information on nicotine is missing in 1 patient]. During their participation in the study, several patients needed sedatives and anxiolytics. Benzodiazepines (BZDs) were administered to 21 of the 50 patients during plasma and CSF samples. At the time of sampling, 58% of the patients (n=29 of 50) were taking antipsychotic medications. For these individuals, the highest number of days on antipsychotic therapy was 25, and the mean number of days was 14.2 ± 2.2 (mean ± s.e.m, n=14). The remaining 21 patients had not used antipsychotics before to or during plasma and CSF collection. The following antipsychotics were used by patients: olanzapine, aripiprazole, risperidone, quetiapine, paroxetine, or haloperidol. Patients' close relatives supplied knowledge available on the duration of untreated psychosis (DUP).

Healthy controls

The study enlisted 47 healthy individuals through advertising. A physical examination, brain magnetic resonance imaging (MRI), blood and urine collection, and other procedures were done to assess the healthy control participants. The Mini International Neuropsychiatric Interview was used to screen subjects for past psychiatric illness. The study excluded participants who used illicit substances, were not on medicine, or had first-degree relatives with mental disorder. During the study, any current drug usage was checked by Alcohol Use Disorders Identification Tests/Drug Use Disorders Identification Tests. An expert neuroradiologist at Karolinska University Hospital, Solna reviewed the MRI data at the MR Centre, and just one participant had a structural anomaly on the MRI. This individual exhibited early demyelination, but it was inadequate for a multiple sclerosis diagnosis because no neurological symptoms had earlier been reported, and a clinical neurological test indicated normal results. Lumbar puncture and cognitive tests for healthy controls were performed within 37.4 ± 5.7 days (mean ± s.e.m.).

CSF collection

For cerebrospinal fluid (CSF) collection, standard lumbar puncture methods were used. A disposable atraumatic needle (22G Sprotte, Pajunk GmbH Medizintechnologie, Geisingen, Germany) was placed at the L 4-5 level in all patients who were in the right decubitus position. CSF (18 ml) was allowed to drop into a plastic test tube protected from light. CSF supernatant from all patients was separated into 10 aliquots and frozen at -80 °C within 1 hour of collection after centrifugation (rotor 5810R, Eppendorf SE, Hamburg, Germany) at 3500 rpm (1438x g) for 10 minutes to separate cells and supernatant. The lumbar puncture was performed on the majority of individuals (n=76; 33 FEP patients and 43 healthy controls) between 07:45 and 10:00 h following a night's sleep. Morning sampling was not achievable for the rest FEP patients (n=17) due to clinical routines. To account for this confounding effect, four healthy individuals were likewise lumbar punctured at the same time window (that is, 10:30 and 13:15 h). Participants in the research were asked not to participate in physical activity for the prior eight hours, although their activity could not be monitored.

sFIDA analysis

CSF samples were provided as a blinded panel to exclude any human bias. Due to the high number of assay points, the measurements were performed on three 384 well microtiter plates, and each sample was applied in quadruplicate. The assay surface was imaged via TIRFM as described above and for analysis, a cutoff of 0.005% of blank control values was used to determine the sFIDA readouts. For assay calibration and as positive control, dilution series (see above) of DISC1 SiNaPs and synthetic DISC1 aggregates, respectively, were applied on each plate. However, since the sFIDA readouts of DISC1 aggregates in human CSF samples were close to buffer control readouts, not enough calibration points of SiNaPs were available to obtain a robust calibration and consequently only the non-calibrated sFIDA readout could be provided for the CSF samples.

*Normalization of sFIDA readouts and statistics*

Because unblinding of the human CSF samples revealed an uneven distribution of healthy controls and FEP samples of all three plates (plate 1: 8 HC, plate 2: 12 HC, plate 3: 27 HC), we normalized the sFIDA readouts based on HC samples to eliminate bias from the statistical results. To this end, the sFIDA readouts of each plate were multiplied by an individual normalization factor (Eq. 4).

$$(4)\quad \text{normalization factor}_{\text{x}} = \frac{\text{mean}_{\text{ HC}} \text{ all plates}}{\text{mean}_{\text{ HC}} \text{ plate x}}$$

Therefore, all sFIDA readouts of plate 1 were multiplied by 0.806, those of plate 2 by 0.980 and those of plate 3 by 1.087 (Supplementary Tab. 1).

Next, we tested the normalized sFIDA readouts for normal distribution (Shapiro-Wilk, Lilliefors, Kolmogorov-Smirnov, Anderson-Darling). Since the data mainly showed non-normal distributions (p-value < 0.05, Supplementary Tab. 4) statistical analysis was performed using non-parametric tests like Mann-Whitney U test or Spearman's correlation. To reveal possible correlations between normalized sFIDA readouts and age or BMI, respectively, we used non-parametric Spearman correlation, both for all 97 samples and for the individual cohorts. In addition, we investigated whether age, smoking, or medication had an effect on the determined sFIDA readouts. To this end, we classified the normalized sFIDA readouts based on the clinical cohorts (HC and FEP) and tested for significant differences using two-sided Mann Whitney U test. To examine whether the levels of DISC1 aggregates in CSF change over time, we compared the normalized sFIDA readouts at baseline with those at the follow-up time point using Wilcoxon rank sum test. Since longitudinal samples were not available for all participants, only those with baseline and follow-up samples were included in the analysis (HC n=15; FEP n=21).

**Supplementary information**

Supplementary Figures

Supplementary Fig. 1. Scheme of the sFIDA assay for the quantitation of DISC1 aggregates...13

Supplementary Fig. 2. sFIDA readout of DISC1 SiNaPs (a) and synthetic DISC1 aggregates (b) diluted in The Blocking Solution...14

Supplementary Fig. 3. sFIDA readouts of inter-assay (a) and inter-machine (b) testing to determine assay comparability. ...15

Supplementary Fig. 4. Assessment of assay selectivity...16

Supplementary Fig. 5. Changes of DISC1 aggregate levels in CSF samples over time. ...17

Supplementary Tables

Supplementary Tab. 1. Normalization factors for individual plates. ...18

Supplementary Tab. 2. Results of different tests on normal distribution of the normalized sFIDA readouts...19

Supplementary Tab. 3. Demographic information on CSF samples. ...20

Supplementary Tab. 4. Spearman coefficient of correlation values for analysis between normalized sFIDA readouts of DISC1 aggregates and age and BMI. ...21

Supplementary Tab. 5. Results of two-sided Mann Whitney U test for analysis of possible influences of sex, smoking, and medication on levels of aggregated DISC1 in CSF samples. ...22

Supplementary Tab. 6. p-values of Mann-Whitney U test for comparison of observed sFIDA readouts of healthy controls and FEP group or subgroup according to their initial preliminary diagnosis. ...23

Supplementary Tab. 7. p-values of Mann-Whitney U test for comparison of observed sFIDA readouts of healthy controls and first-episode psychosis group or subgroup according to their final clinical diagnosis after 18 months. ........................................................................................24

Supplementary Tab. 8. Correlations of DISC1 aggregate levels with symptom levels and cognitive function……………………………………………………………………………………………26

### Supplementary Figures

**Supplementary Fig. 1. Scheme of the sFIDA assay for the quantitation of DISC1 aggregates.**

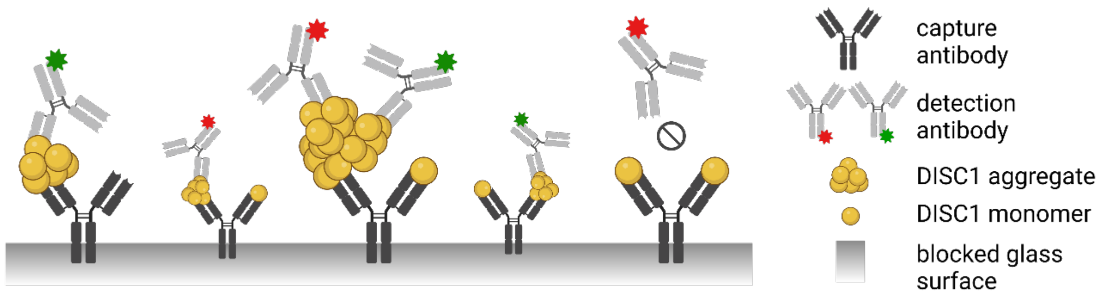


The glass surface of a microtiter plate is coated with 14F2 antibodies directed against linear epitopes of DISC1 (aa 747-768). Afterwards, human CSF sample is incubated on assay surface, with both monomeric and aggregated DISC1 immobilized by the capture antibodies. However, the fluorescently labelled detection antibodies (14F2 CF633 and 14F2 CF488A) can only detect aggregated DISC1 species because all assay antibodies bind to the same epitope. In case of monomeric DISC1, the epitope is occupied by the capture antibody, preventing binding of the probe antibody. Finally, the assay surface is imaged by dual-color TIRF microscopy and only single particles with co-localized fluorescence signals on the well surface are counted by image-data analysis. *Created with BioRender.com.*

**Supplementary Fig. 2. sFIDA readout of DISC1 SiNaPs (a) and synthetic DISC1 aggregates (b) diluted in The Blocking Solution**

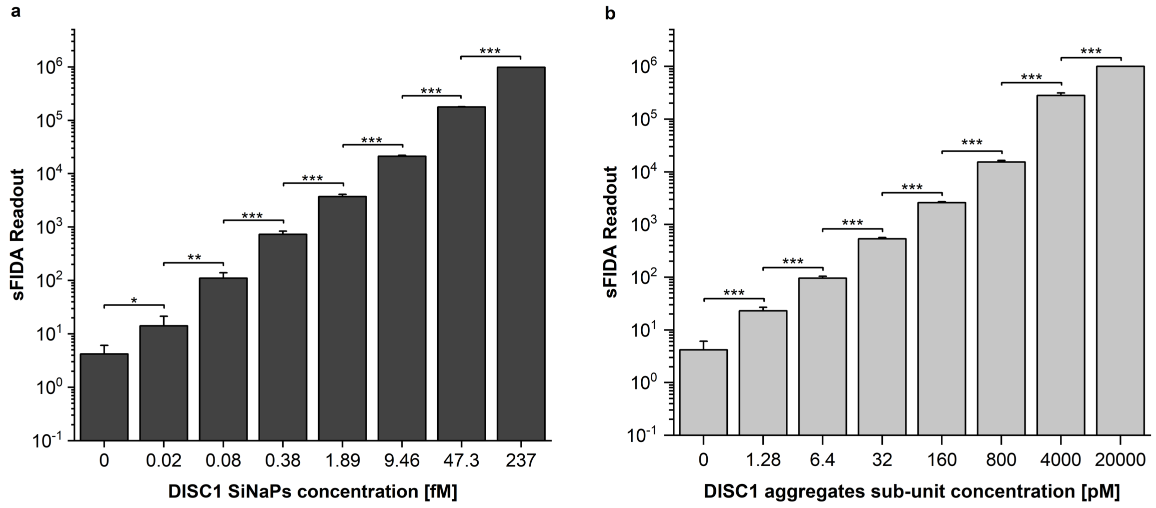


Columns and error bars represent the calculated arithmetic mean and standard deviations from a fourfold determination of DISC1 SiNaPs and synthetic DISC1 aggregates, respectively. The blank control was determined 8-fold. To decrease background noise, blank control-based cutoffs were defined as the signal intensity exceeding 0.05% of total pixels. Significant difference between each concentration compared to the next lower one was determined by one-sided Mann-Whitney U test (confidence interval 0.05, * $p \leq 0.05$; ** $p \leq 0.01$; *** $p \leq 0.001$). For both targets, a linear correlation between sFIDA readout and the applied concentration was observed, indicated by a Pearson's coefficient of correlation r=0.9997 for DISC1 SiNaPs and r=0.9958 for DISC1 aggregates. For calculation of the calibration curve, all DISC1 SiNaPs concentrations were included, since all of them were in the linear measurement range and significantly differed from blank control. Using calibration curve (y=3.908*x-13.79, $R^2$=0.9828), LoD and LLoQ were calculated based on blank control readouts to 6.08 fM and 9.54 fM, respectively. In addition, intra-assay variability was determined with a mean CV% of 15.8% for DISC1 SiNaPs and 7.9% for synthetic DISC1 aggregates, respectively.

**Supplementary Fig. 3. sFIDA readouts of inter-assay (a) and inter-machine (b) testing to determine assay comparability.**

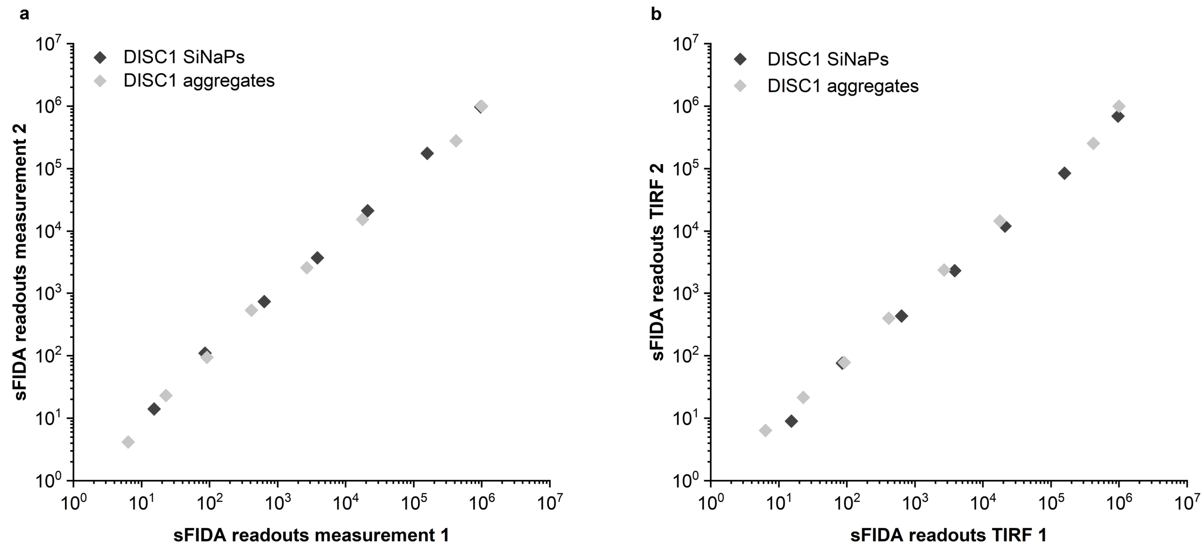


We investigated both inter-assay and inter-machine variability, respectively, of the sFIDA assay for DISC1 SiNaPs and DISC1 aggregates. **(a)** sFIDA readouts of two independent measurements of both targets by the same operator on different days. **(b)** Testing inter-machine variations, the same assay plate was measured on two different TIRF microscopes (TIRF 1 and TIRF 2). The developed assay is able to achieve comparable results within two independent experiments as indicated by an average percent comparability (Eq. 2) of 106.5% for DISC1 SiNaPs and 97.5% for DISC1 aggregate dilution series. In addition, both TIRF microscopes delivered similar results, with a percent comparability of 98.7% for DISC1 SiNaPs and 91.3% for DISC1 aggregate dilution series.

**Supplementary Fig. 4. Assessment of assay selectivity**

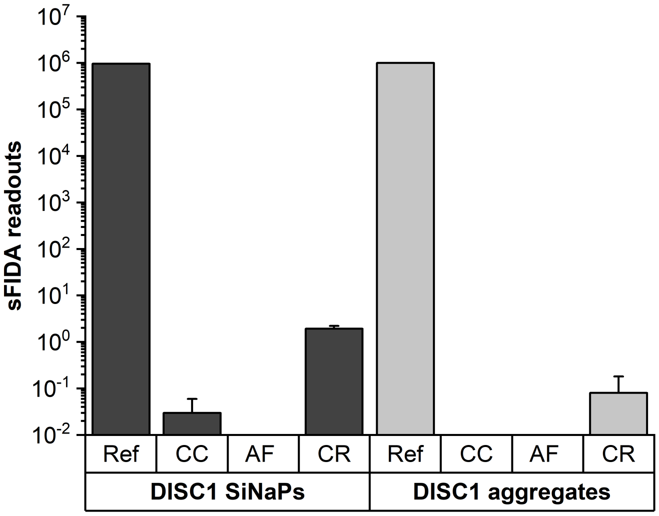


To assess assay selectivity, we applied DISC1 SiNaPs (237 pM, molar particle concentration) and synthetic DISC1 aggregates (20 nM, molar sub-unit concentration) on the standard assay surface (reference, ref) as well as on various assay controls setups (CC: capture control w/o capture antibody; AF: autofluorescence control w/o detection antibodies; CR: cross-reactivity control, detection antibodies directed against amyloid β). Afterwards, observed sFIDA readouts of each assay control were compared to reference values and percent signal reduction was calculated (Eq. 3a and 3b). For both DISC1 SiNaPs and synthetic DISC1 aggregates, the sFIDA readouts were reduced by 99-100% in all controls.

**Supplementary Fig. 5. Changes of DISC1 aggregate levels in CSF samples over time.**

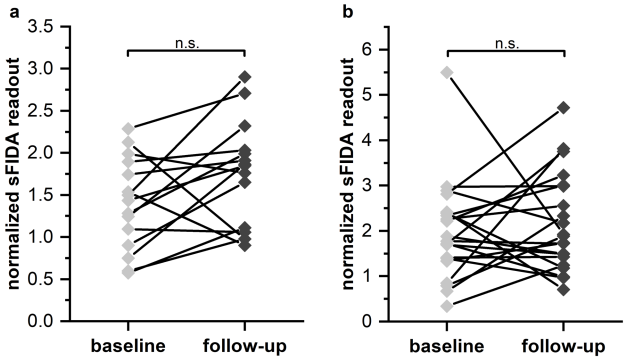


Baseline (inpatient admission) and follow-up levels (re-assessment) of DISC1 aggregate in CSF of 15 healthy controls **(a)** and 21 FEP subjects **(b)** were investigated for significant changes by Wilcoxon rank sum test (confidence interval 0.05). For both, healthy controls and diseased samples, no significant changes in biomarker levels were observed, as indicated by p-value of 0.074 (healthy controls) and p-value 0.487 (FEP samples), respectively.

### Supplementary Tables

**Supplementary Tab. 1. Normalization factors for individual plates.**

| Plate No. | Normalization factor |
|---|---|
| 1 | 0.80649 |
| 2 | 0.97952 |
| 3 | 1.08741 |

**Supplementary Tab. 2. Results of different tests on normal distribution of the normalized sFIDA readouts.**

| | Shapiro-Wilk | Lilliefors | Kolmogorov-Smirnov | Anderson-Darling |
|---|---|---|---|---|
| **p-values** | $5.69*10^{-8}$ | $3.46*10^{-4}$ | 0.068 | $6.41*10^{-7}$ |

p-values < 0.05 indicated non-normal distribution of the data

**Supplementary Tab. 3. Demographic information on CSF samples.**

| | all cohorts | HC | FEP | Kruskal-Wallis ANOVA |
|---|---|---|---|---|
| **Number** | 97 | 47 | 50 | |
| **Female** | 58 (59.8%) | 27 (57.4%) | 31 (62.0%) | 0.05652 |
| **Age [mean ± s.e.m]** | 28.52 ± 0.74 | 26.85 ± 0.78 | 30.08 ± 1.19 | 0.07229 |
| **BMI [mean ± s.e.m]** | 23.84 ± 0.38 | 24.23 ± 0.55 | 23.47 ± 0.53 | 0.44859 |
| **Nicotine yes** | 20 (20.6%) | 7 (14.9%) | 13 (22.8%) | 0.16267 |
| **Medication yes** | 29 (29.9%) | 0 (0%) | 29 (58%) | *$5.52*10^{-10}$* |

Kruskal-Wallis ANOVA: confidence interval 0.05. Significant differences between both cohorts are written in italics.

**Supplementary Tab. 4. Spearman coefficient of correlation values for analysis between normalized sFIDA readouts of DISC1 aggregates and age and BMI.**

| Spearman r | all cohorts | HC | FEP |
|---|---|---|---|
| **Age** | -0.0623 (p: 0.5445) | -0.2399 (p: 0.1043) | -0.0032 (p: 0.9824) |
| **BMI** | -0.0859 (p: 0.4027) | 0.0040 (p: 0.9788) | -0.1660 (p: 0.2494) |

No significant correlation was found

**Supplementary Tab. 5. Results of two-sided Mann Whitney U test for analysis of possible influences of sex, smoking, and medication on levels of aggregated DISC1 in CSF samples.**

| | comparison | HC (p-value) | FEP (p-value) |
|---|---|---|---|
| **sex** | f vs m | 0.8464 | 0.6033 |
| **nicotine** | yes vs no | 0.4550 | 0.2435 |
| **medication** | yes vs no | - | 0.2797 |

Confidence interval 0.05

**Supplementary Tab. 6. p-values of Mann-Whitney U test for comparison of observed sFIDA readouts of healthy controls and FEP group or subgroup according to their initial preliminary diagnosis.**

| **HC vs.** | **p-value** |
|---|---|
| FEP | *0.0257* |
| ATPDU | - |
| DD | 0.2363 |
| HS | - |
| MD | - |
| OS | 0.1778 |
| PS | 0.4008 |
| SAD | - |
| SU | 0.0903 |
| UNO | *0.0048* |

Significant differences (p-values < 0.05) between FEP (sub)group and HC are written in italics. Please note that for the comparison of HC vs group FEP two-sided and for the comparison of HC vs each subgroup one-sided Mann-Whitney U test was performed, respectively. No Mann-Whitney U test was performed for ATPDU, HS, MD and SAD, as only one value was available.

*HC* healthy control, *FEP* first-episode psychosis patients, *ATPDU* Acute and transient psychotic disorder, unspecified (DSM IV 298.8), *DD* Delusional Disorders (DSM IV 297.1), *HS* hebephrenic schizophrenia (DSM IV 295.10), *MD* Major depressive disorder (DSM IV 296.21), *OS* Other Schizophrenia (DSM IV 295.40), *PS* Paranoid schizophrenia (DSM IV 295.30), *SAD* Schizoaffective disorder (DSM IV 295.70), *SU* Schizophrenia, unspecified (DSM IV 295.9), *UNO* Unspecified nonorganic psychosis (DSM IV 298.9).

**Supplementary Tab. 7. p-values of Mann-Whitney U test for comparison of observed sFIDA readouts of healthy controls and first-episode psychosis group or subgroup according to their final clinical diagnosis after 18 months.**

| **HC vs.** | **p-value** |
|---|---|
| FEP | *0.0257* |
| ATPDU | - |
| DD | 0.7094 |
| HS | 0.1181 |
| MD | - |
| OS | - |
| PS | 0.2911 |
| RS | - |
| SAD | *0.0012* |
| SU | *0.0235* |
| UNO | 0.1953 |

Significant differences (p-values < 0.05) between FEP (sub)group and HC are written in italics. Please note that for the comparison of HC vs group FEP two-sided and for the comparison of HC vs each subgroup one-sided Mann-Whitney U test was performed, respectively. No Mann-Whitney U test was performed for ATPDU, MD, OS and RS as only one value was available.

*HC* healthy control, *FEP* first-episode psychosis, *ATPDU* Acute and transient psychotic disorder, unspecified (DSM IV 298.8), *DD* Delusional Disorders (DSM IV 297.1), *HS* hebephrenic schizophrenia (DSM IV 295.10), *MD* Major depressive disorder (DSM IV 296.21), *OS* Other Schizophrenia (DSM IV 295.40), *PS* Paranoid schizophrenia (DSM IV 295.30), *RS* Residual Schizophrenia (DSM IV 295.60), *SAD* Schizoaffective disorder (DSM IV 295.70), *SU*

Schizophrenia, unspecified (DSM IV 295.9), *UNO* Unspecified nonorganic psychosis (DSM IV 298.9).

**Supplementary Tab. 8. Correlations of DISC1 aggregate levels with symptom levels and cognitive function.**

| Baseline | PANSS Positive | PANSS Negative | PANSS General | PANSS Total | GAF-Symp | GAF- Func | CGI |
|---|---|---|---|---|---|---|---|
| *Spearman Correlation* | -0,08 | -0,49 | -0,01 | -0,2 | 0,07 | 0,19 | -0,1 |
| *p-value* | 0,72 | 0,023 | 0,95 | 0,37 | 0,75 | 0,4 | 0,66 |
| Adjusted p-value | | | | | | | |

| Follow-up | PANSS Positive | PANSS Negative | PANSS General | PANSS Total | GAF-Symp | GAF- Func | CGI |
|---|---|---|---|---|---|---|---|
| *Spearman Correlation* | 0,08 | 0,02 | 0,34 | 0,21 | -0,17 | -0,02 | -0,08 |
| *p-value* | 0,72 | 0,93 | 0,15 | 0,37 | 0,49 | 0,93 | 0,73 |
| Adjusted p-value | | | | | | | |

| Baseline | TMT | BACS_SC | HVLT_R | WMS_II_SS | LNS | NAB_MAZES | BVMT_R | Fluency | MSCEIT_ME | CPT_IP |
|---|---|---|---|---|---|---|---|---|---|---|
| *Spearman Correlation* | -0,05 | 0,56 | 0,29 | 0,09 | 0,29 | 0,31 | 0,29 | 0,33 | -0,13 | 0,27 |
| *p-value* | 0,81 | 0,008 | 0,21 | 0,69 | 0,21 | 0,17 | 0,19 | 0,14 | 0,58 | 0,24 |
| Adjusted p-value | | | | | | | | | | |

| Follow-up | TMT | BACS_SC | HVLT_R | WMS_II_SS | LNS | NAB_MAZES | BVMT_R | Fluency | MSCEIT_ME | CPT_IP |
|---|---|---|---|---|---|---|---|---|---|---|
| *Spearman Correlation* | 0,23 | 0,06 | 0,08 | 0,31 | 0,11 | 0,07 | 0,13 | 0,04 | 0,09 | 0,23 |
| *p-value* | 0,34 | 0,81 | 0,75 | 0,18 | 0,64 | 0,78 | 0,58 | 0,86 | 0,69 | 0,35 |
| Adjusted p-value | | | | | | | | | | |

No significant correlations (after false discovery rate (FDR) corrections, not included in the table) between DISC1 levels in CSF and the following variables [6-9] were observed: PANSS (positive and negative syndrome scale), GAF-Symp (global assessment of functioning), GAF-Func, CGI (clinical global impression), TMT (trail making test), BACS_SC (brief assessment of cognition in schizophrenia-symbol coding test), HVLT_R (Hopkins verbal learning test—revised), WMS_II_SS_R (Wechsler memory scale III spatial span test), Fluency, MSCEIT_ME (Mayer-Salovey-Caruso emotional intelligence test's managing emotions component), CPT_IP (continuous performance test—identical pairs).

**References**


1. Hülsemann M*, et al.* Biofunctionalized Silica Nanoparticles: Standards in Amyloid-β Oligomer-Based Diagnosis of Alzheimer's Disease. . *J Alzheimers Dis.* **54**, 79-88 (2016).

2. Blömeke L*, et al.* Quantitative detection of α-Synuclein and Tau oligomers and other aggregates by digital single particle counting. . *NPJ Parkinsons Dis.* **8(1):68**, 68 (2022).

3. Ottis, P.*, et al.* Convergence of two independent mental disease genes on the protein level: recruitment of dysbindin to cell invasive DISC1 aggresomes. *Biol Psychiatry* **70**, 604-610 (2011).

4. Leliveld, S.R.*, et al.* Oligomer assembly of the C-terminal DISC1 domain (640-854) is controlled by self-association motifs and disease-associated polymorphism S704C. *Biochemistry* **48**, 7746-7755 (2009).

5. Kravchenko K*, et al.* Analysis of anticoagulants for blood-based quantitation of amyloid β oligomers in the sFIDA assay. . *Biol. Chem.* **398**, 465-475 (2017).

6. Kay, S.R., Fiszbein, A. & Opler, L.A. The positive and negative syndrome scale (PANSS) for schizophrenia. *Schizophr Bull* **13**, 261-276 (1987).

7. Aas, I.H. Guidelines for rating Global Assessment of Functioning (GAF). *Ann Gen Psychiatry* **10**, 2 (2011).

8. Busner, J. & Targum, S.D. The clinical global impressions scale: applying a research tool in clinical practice. *Psychiatry (Edgmont)* **4**, 28-37 (2007).

9. Nuechterlein, K.H.*, et al.* The MATRICS Consensus Cognitive Battery, part 1: test selection, reliability, and validity. *Am J Psychiatry* **165**, 203-213 (2008).
